# Multivariate genetic of 2.2 million individuals demonstrate genetic influences on substance use disorders operate via behavioral disinhibition and substance-specific risk

**DOI:** 10.1101/2024.11.26.24318011

**Authors:** Holly E. Poore, Chris Chatzinakos, Brittany Leger, Jean Gonzalez, Travis T. Mallard, Sandra Sanchez-Roige, Fazil Aliev, Alexander Hatoum, COGA Collaborators, Irwin D. Waldman, Abraham A. Palmer, K. Paige Harden, Danielle M. Dick, Peter B. Barr

**Affiliations:** Department of Psychiatry, Robert Wood Johnson Medical School, Rutgers University; Department of Psychiatry and Behavioral Science, SUNY Downstate Health Sciences University; Department of Psychiatry, University of California San Diego; Institute for Genomic Medicine, University of California San Diego; Center for Precision Psychiatry, Department of Psychiatry, Massachusetts General Hospital; Department of Psychiatry, Harvard Medical School; Department of Medicine, Vanderbilt University Medical Center; Department of Psychiatry, Washington University School of Medicine; Department of Psychology, Emory University; Department of Psychology, University of Texas at Austin; Population Research Center, University of Texas at Austin

## Abstract

Ongoing efforts to identify genes involved in substance use disorders (SUDs) often focus on individual disorders despite high rates of co-occurrence with each other and other externalizing traits. Here, we investigate whether incorporating data on other externalizing traits can boost power to detect without sacrificing specificity of SUD genetic signal. We used multivariate genomic analyses and downstream biological annotation and genetic association analyses to explore this question. We found that joint analysis of SUDs and other externalizing traits resulted in increased insights into the neurobiology of broad and substance-specific SUD risk. We found no evidence of loss of specificity for SUD genetic signal but note improvements in our ability to characterize the neurobiology of broad and substance-specific SUD genetic effects. Our findings suggest that genetic risk for SUDs operates largely via pathways shared with other behaviors characterized by behavioral disinhibition, with additional substance-specific risk, and that modeling this shared disposition improves gene discovery.

---

Substance use disorders (SUDs) are common^1^ and associated with significant cost to the individuals, their families, and society^2^. They are also significantly heritable (∼40-60%)^3-5^, leading to several on-going efforts to identify genes that confer risk for SUDs. Most of these studies focus on specific drugs (e.g., smoking^6^ or alcohol separately) However, SUDs frequently co-occur, and twin studies indicate that this overlap is largely due to shared genetic influences. This shared genetic influence, which broadly impacts SUD risk, accounts for up to 74-80% of the genetic influences on alcohol use disorders and 62-74% of genetic influences on other substance use disorders^5,7^, with the remaining genetic risk being substance specific. More recently, multivariate genome-wide association studies (GWAS) have been applied to SUDs, providing further evidence that most genomic risk is shared^8-11^.

We also know from epidemiological and twin studies that SUDs share phenotypic and genetic influences with other outcomes characterized by behavioral disinhibition; SUDs load together with childhood conduct disorders, adult antisocial behavior, and personality traits on a common underlying genetic factor^12,13^. In twin studies, this underlying latent factor is highly heritable (∼80%), moreso than any of the disorders or traits studied individually. In the psychological literature, this spectrum of behaviors and disorders, including SUDs, is typically referred to as the externalizing spectrum^14,15^.

Despite this strong evidence that SUDs and other traits related to behavioral disinhibition can be modeled with a common dimension of phenotypic and genotypic variation, most gene identification efforts for SUDs study each disorder in isolation or focus on a single dimension of SUD risk^8,10^. The question continues to arise: is the genetic variation captured by externalizing distinct from genetic risk for SUDs or are the genetic influences on SUDs largely those that impact multiple externalizing conditions? The answer to this question can be used to inform the best way to study the etiology of SUDs. If genetic variance between SUDs and externalizing traits is mostly shared, we can use well-powered externalizing GWAS results to boost signal for SUDs. On the other hand, if genetic variation is sufficiently distinct, we risk masking SUD-specific genetic signal in a joint analysis.

As a first step in addressing this question, our group recently published a study using Genomic SEM^16^ to test alternative models of the relationship between four SUDs, for which large GWAS are now available, and six other externalizing traits used in a previous GWAS of externalizing^17^. The two best fitting models were (1) a common factor in which all SUDs and other externalizing traits loaded onto a single factor, indicating that genetic risk for SUDs is largely indistinguishable from broad externalizing risk, and (2) a two-factor model in which the factor representing shared SUD risk was correlated with a factor representing behavioral disinhibition, suggesting that there was substantial overlap between SUDs and other externalizing traits, but some specificity for SUDs remained. Either way, results from this study suggested that the genetic influences on these two sets of phenotypes were highly correlated. Although both models fit well, the correlation between the two dimensions in the second model was ∼.90, suggesting they share the majority of their genetic influences.

In the current study, we carry forward both models to conduct multivariate genomic analyses from data on > 2.2 million individuals on these three factors. We use Genomic SEM and a series of downstream biological annotation and genetic association analyses to answer three main questions: 1) does joint analysis of behavioral disinhibition traits and SUDs result in increased power and/or decreased specificity to detect genetic effects for SUDs? 2) what can we learn about the neurobiology specific to each SUD (i.e., after accounting for what SUDs share with other externalizing indicators)? and 3) what do we learn about risk for addiction broadly when we analyze these traits together?

## Results

### Multivariate GWAS

We used Genomic SEM^16^ to perform multivariate GWAS on latent factors that posit different ways that genetic influences impact SUDs and correspond to two models previously estimated by this group^18^ (Supplementary Table 1a and b). The first model hypothesizes that a shared set of genes influences SUDs along with other externalizing phenotypes, such as attention deficit hyperactivity disorder (ADHD) and substance initiation (see below for further discussion of each phenotype). This shared genetic dimension in shown in Box A of Figure 1. This model also facilitates examination of the genetic influences on each SUD after accounting for what it shares with other SUDs and externalizing traits (Boxes B-E in Figure 1). These influences reflect residual, substance-specific genetic effects after accounting for what is shared with each of the other externalizing indicators, hereafter referred to as “residual-SUD”). The second model hypothesizes that separate, but correlated, dimensions of genetic risk influence SUDs and other externalizing traits, here referred to as behavioral disinhibition (Boxes F and G in Figure 1). In our previous work^18^, both models emerged as viable candidates; thus, in this study, we carry both forward into the multivariate GWAS analyses to compare their ability to facilitate gene identification for SUDs.

**Figure 1.**
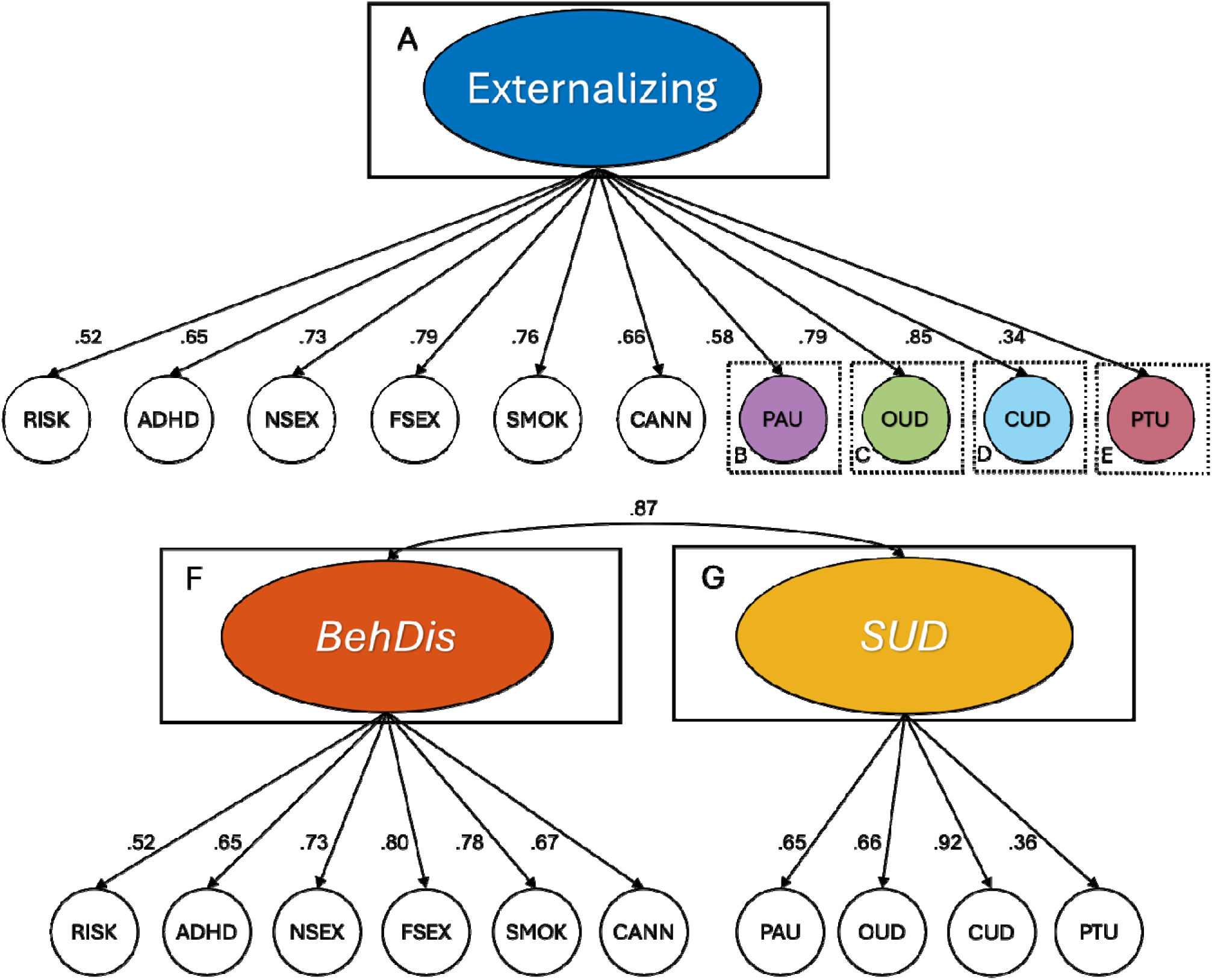
Path diagrams of models used in the current analyses. Box A represents the broad Externalizing factor onto which all behavioral disinhibition and SUD phenotypes load. Boxes B-E represent residual SUD phenotypes. Boxes F and G represent narrower factors reflecting Behavioral Disinhibition and SUD phenotypes, respectively. Single headed arrows indicate factor loadings whereas the double headed arrow indicates a correlation between the two factors. RISK = risk taking, ADHD = attention deficit hyperactivity disorder, NSEX = number of sexual partners, FSEX = age at first sexual intercourse, SMOK = smoking initiation, CANN = cannabis initiation, PAU = problematic alcohol use, OUD = opioid use disorder, CUD = cannabis use disorder, PTU = problematic tobacco use.

As this study is a direct extension of and requires comparison to results from our previous work ^17,18^, we retain the GWAS used in those studies as indicators in the current models. These indicators include: ADHD^19^, cannabis initiation (CANN)^20^, smoking initiation (SMOK)^21^, age at first sexual intercourse (FSEX)^22^, number of sexual partners (NSEX)^22^, risk taking propensity (RISK)^22^, problematic alcohol use (PAU)^23^, problematic tobacco use (PTU)^9^, opioid use disorder (OUD)^24^, and cannabis use disorder (CUD)^25^. Due to concerns about the use of FSEX as an externalizing indicator^26,27^ we rab sensitivity analyses leaving out this indicator, which indicated that it did not exert undue influence on the composition of the factor (see Supplementary Text and Supplementary Table 2 for more detail). All GWAS included only individuals whose genomes were most similar to those from reference panels sampled from Europe (hereafter referred to as “EUR”) with a total sample size of 2,219,357 unique individuals (Supplementary Table 3). After merging across the ten sets of summary statistics, 5,963,905 SNPs were available for analysis.

We identified 708 loci in the single factor Externalizing model, in which SUDs were modeled as part of the externalizing spectrum (Supplementary Table 1a). In the two-factor model, we identified 631 and 48 genomic risk loci (679 total) for the Behavioral Disinhibition and SUD factors, respectively (Figure 2; Supplementary Table 4; see Supplementary Text for additional details). Of the 708 Externalizing loci and their correlates within LD regions (*r^2^* > .1), 187 (26%) were not identified in the previous Externalizing^17^ or Addiction Risk^8^ GWAS and 403 (57%) were not previously associated with a substance use trait in the GWAS literature (as reported in the NHGRI-EBI GWAS Catalog^29^ version e114_r2025-06-27). Novel variants include rs11692435 mapped to *ACTR1B*, which is involved in a variety of cellular functions and has been previously implicated in alcoholic gastritis, alcohol consumption, and smoking initiation and rs10145520, mapped to *RAB2B,* which is involved in GTP-binding and has been previously implicated in smoking initiation and educational attainment.

**Figure 2.**
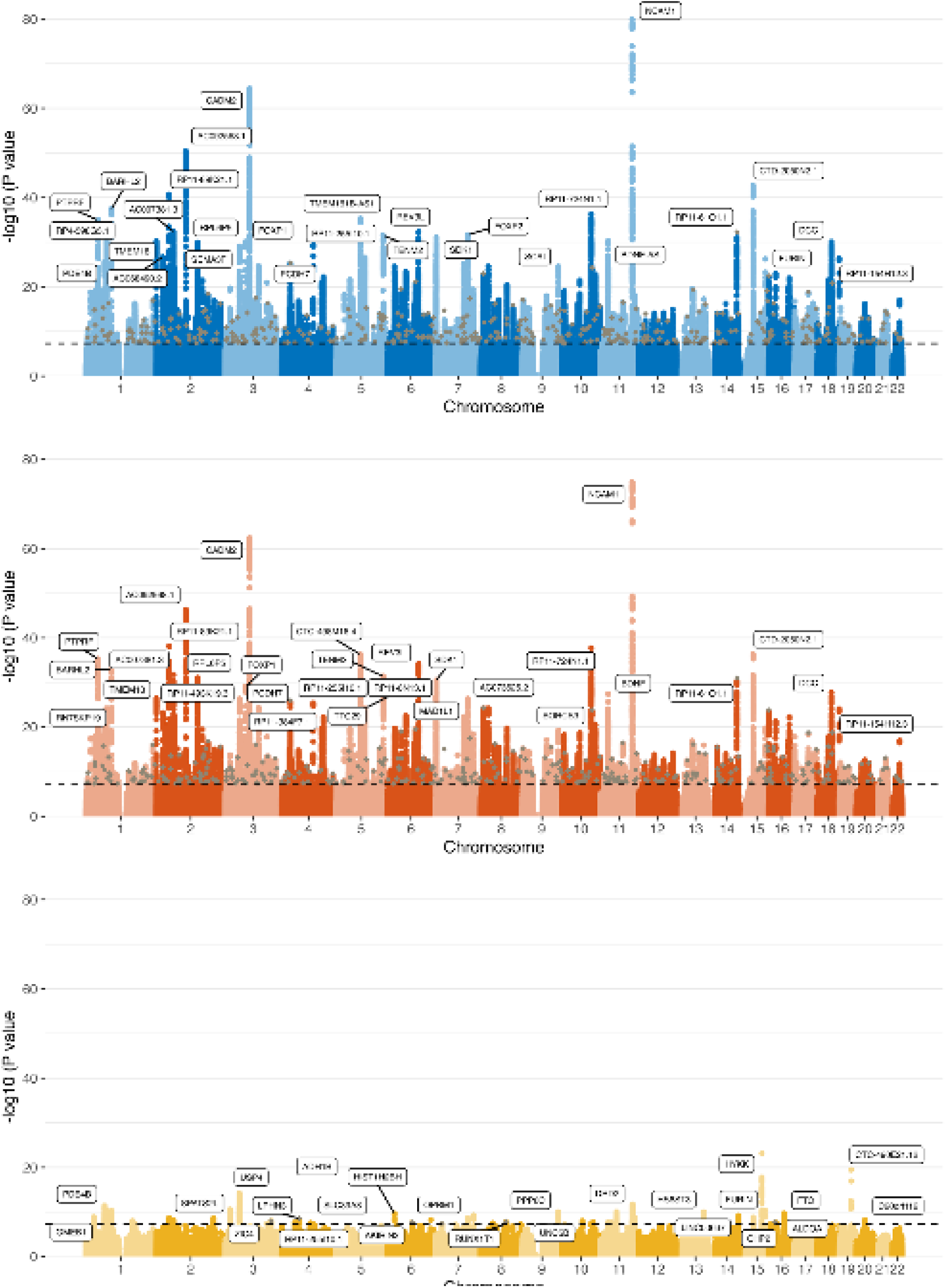
Manhattan plots of (top to bottom) Externalizing, Behavioral Disinhibition, and SUD. Brown points represent novel SUD loci (loci not previously associated with a substance use trait). Top loci are mapped to the nearest gene using ANNOVAR^28^ annotation.

We identified 14, 23, and 1 genomic risk loci for the residual-PAU, PTU, and CUD phenotypes, respectively, whereas the residual-OUD GWAS did not identify any genome-wide significant hits (Supplementary Table 5). Many of these loci were mapped to genes involved in pharmacokinetic (e.g., metabolism) or pharmacodynamic (e.g., drug targets) processes for specific substances. For example, residual-PAU loci were mapped to genes in the alcohol dehydrogenase family (*ADH1B, ADH4, ADH5, ADH6*), which is involved in metabolism of alcohol, and loci for residual-PTU were mapped to genes in the nicotinic acetylcholine receptor family of proteins (*CHRNA3, CHRNA5, CHRNA6, CHRNB3, CHRNB4)*.

### Biological annotation

We used four methods to identify genes associated with the three latent genomic factors: multi-marker analysis of genomic annotation (MAGMA; version 1.08; Supplementary Tables 6 and 7)^30^, MetaXcan^31^ (Supplementary Tables 8), summary-based Mendelian randomization (SMR; Supplementary Tables 9)^32^, and Hi-C-coupled MAGMA (H-MAGMA; Supplementary Tables 10 and 11)^33^. MAGMA tissue expression analyses are reported in the Supplementary Text and Supplementary Tables 12 and 13.

#### Identifying genes for latent genomic factors

To account for differences in gene-based mapping methods, we further evaluated the intersection of genes that were identified in all four gene-based analyses. The intersection of genes identified by these four methods resulted in 113 genes for Externalizing, and 97 and 4 (101 total) genes for Behavioral Disinhibition and SUDs, respectively (Figure 3a; Supplementary Table 14). We next identified genes that were unique to each factor (i.e. associated with one and not the other two factors) and found 37 unique genes for Externalizing and 21 for Behavioral Disinhibition and 3 for SUDs in the two-factor model (Figure 3b). Of the 37 genes unique Externalizing genes, 30 (81%) have been previously associated with a substance use phenotype (e.g., initiation, consumption, or use disorder),^29^ including *SMIM19*, which has been associated with nicotine dependence, and *KLHL29*, which has been associated with cannabis, nicotine, and alcohol use. Of the three unique SUD genes, *PPP6C* has been associated with CUD, AUD, and OUD.

**Figure 3.**
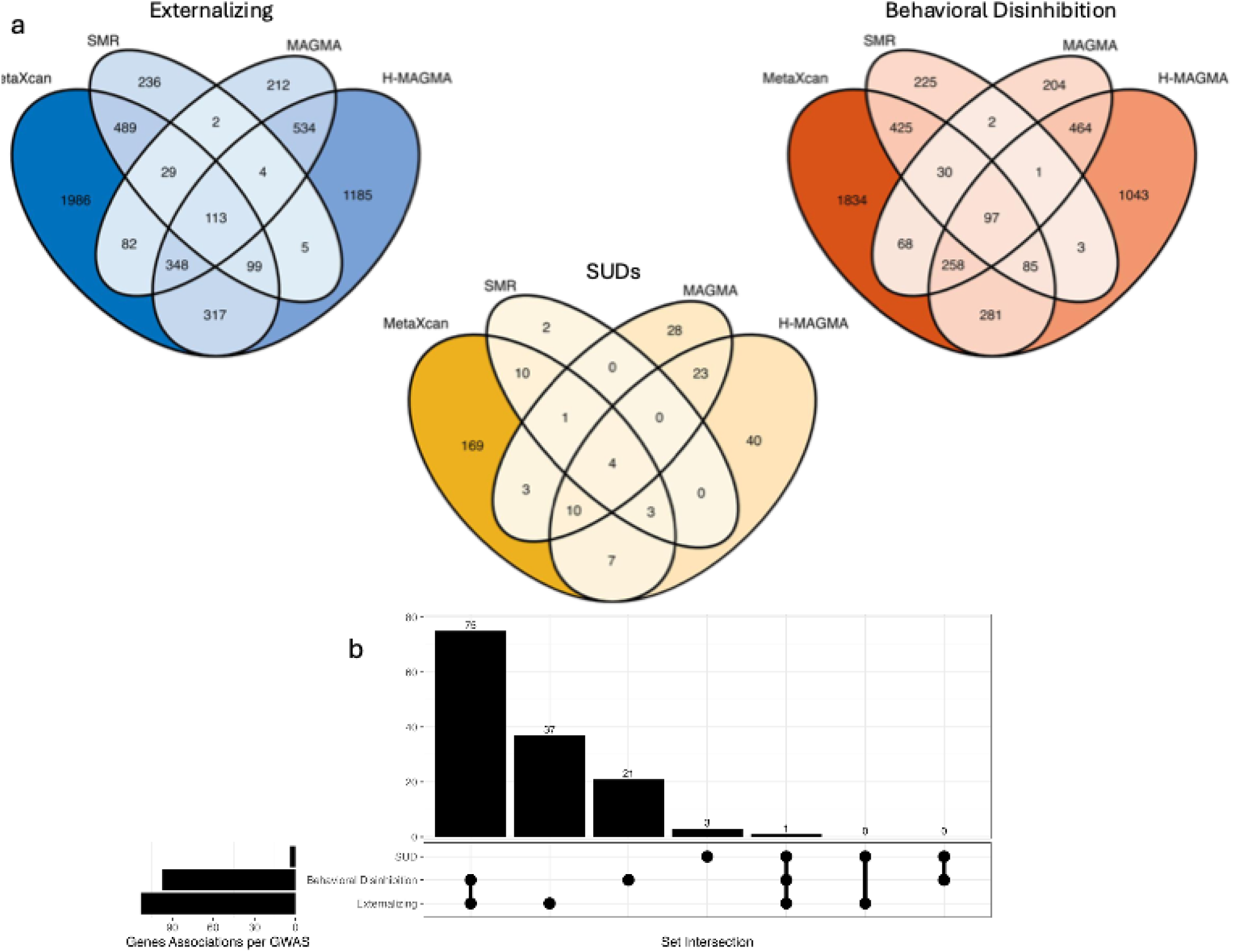
(a) Venn diagrams showing the overlap of genes identified by the MetaXcan, SMR, MAGMA, and H-MAGMA analyses for Externalizing (blue), Behavioral Disinhibition (orange), and SUD (yellow). (b) upset plot showing intersecting sets of high confidence genes (those identified by all three gene-based methods) across Externalizing, Behavioral Disinhibition, and SUD.

#### Identifying residual SUD genetic influences

Only genes for residual-PAU and PTU were identified in multiple methods (and only using MAGMA and H-MAGMA methods), resulting in 21 and 25 genes, respectively (Supplementary Table 15). We saw some substance-specificity in the genes associated with the residual SUD phenotypes. Most genes identified for the residual SUDs were previously associated with a substance use phenotype and many were associated *only* with a phenotype for that specific substance. For example, of the 21 genes associated with residual-PAU, the 13 genes that were previously associated with a substance use trait were *only* associated with an alcohol and not with other substances (excluding multivariate analyses of multiple substances).

#### Network analysis

We employed gene-network analysis^34,35^ for each factor to identify genes that are part of the implicated biological networks, even if they are not directly identified by the GWAS (Figure 4a; see Supplemental Text for more detail). The Externalizing and Behavioral Disinhibition networks were similar to one another, whereas the SUD network shared few genes, likely due to the limited power of the SUD gene set. We then used hierarchical clustering^36^ and gene ontology^37,38^ enrichment to identify the gene communities within these networks (Supplementary Table 16). We found that the Externalizing network contains gene communities that function in post-synaptic protein localization, organization, and signaling, as well as vesicle-mediated transport, and cytoskeleton-dependent transport (Figure 4b). We further validated our findings using annotations in the GWAS catalog. We found that the Externalizing network was significantly enriched for genes identified in psychiatric disorders (p = 2.2×10^-7^), substance-related disorders (p = 2.5×10^-10^), and substance abuse (p = 4.5×10^-14;^ Supplementary Table 17). The Behavioral Disinhibition network showed a similar pattern of enrichment, whereas the SUD network was not significantly enriched for genes identified in any traits in the GWAS catalog.

**Figure 4:**
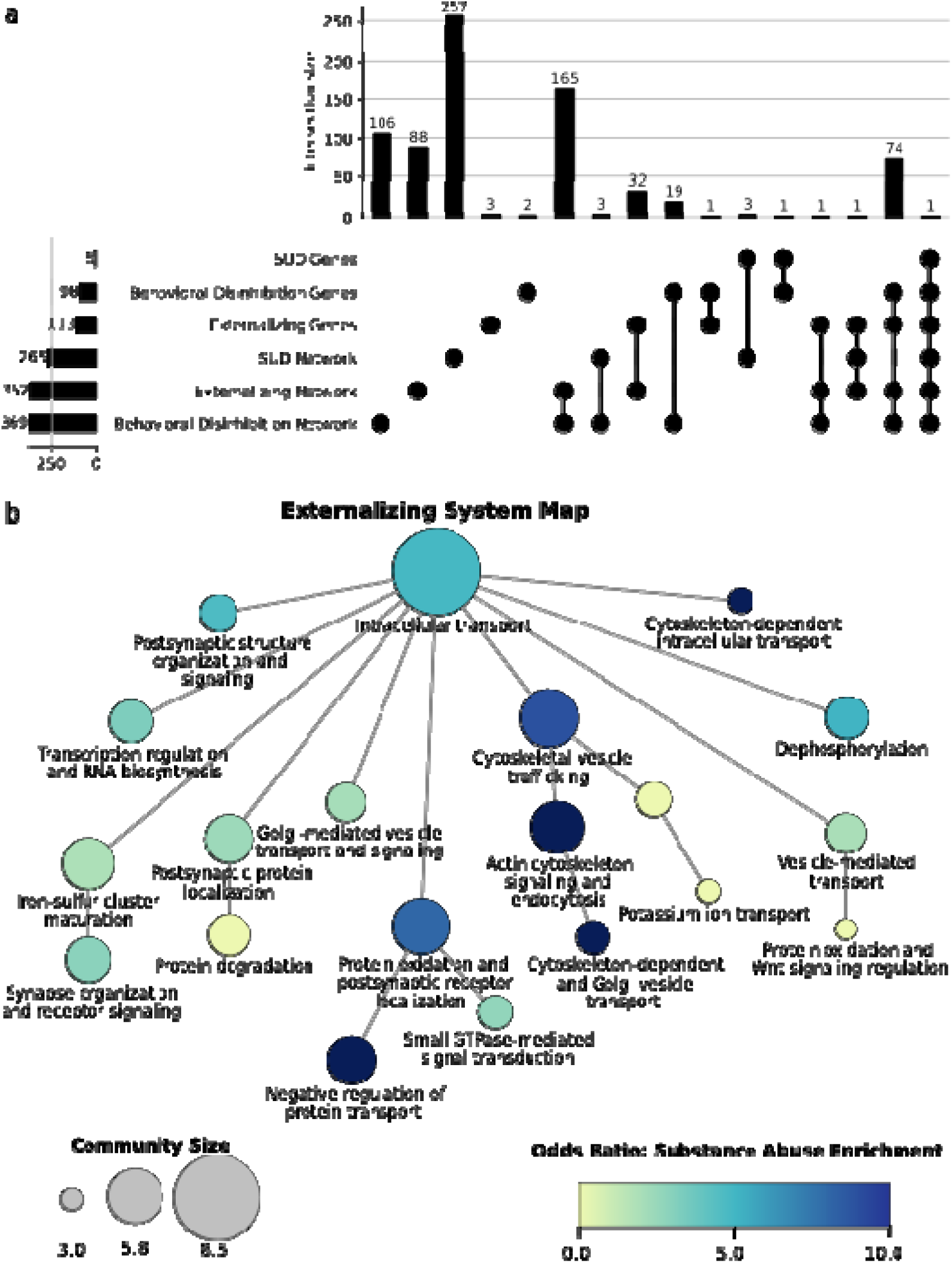
Network Annotation. (a) Upset plot showing the overlap in gene s identified from gene mapping algorithms (see Figure 3a), and network analysis. (b) System map for the Externalizing network. Community labels represent aggregated significant gene ontology enrichment labels, and node size indicates log-transformed community size, measured by number of genes. Color indicates the odds ratio of the enrichment of that community for genes annotated in the GWAS catalog for substance abuse. Only labelled communities and those connecting labeled communities are shown.

#### Drug repurposing

We next mapped genes associated with factors and residual SUDs to the Drug-Gene-Interaction Database to identify druggable targets, with a particular emphasis on drugs with FDA approval for use in medication assisted treatment (MAT) of SUDs (Supplementary Table 18). For the factors, we used genes identified in at least three of the four gene-based methods. Given their lower power and the exploratory nature of these analyses, we used genes identified in any gene-based method for the residual SUD effects. We identified 118 druggable targets resulting in 1,024 FDA approved drug-gene interactions for Externalizing. Identified drugs included those used in MAT, including naltrexone, methadone hydrochloride, disulfiram, varenicline, baclofen, and acamprosate calcium. Genes identified for Behavioral Disinhibition and SUDs resulted in 96 and three druggable targets and 704 and 11 drug-gene interactions, respectively. Drugs used in MAT were also identified for each factor, but fewer types of drugs and at a lower rate compared with Externalizing. We found 16 and 28 druggable targets and 181 and 231 drug-gene interactions for residual PAU and PTU, respectively (Supplementary Table 19). We found that the associations for residual-PAU were specific; the only MAT drugs identified, disulfiram and naltrexone, are used to treat problematic alcohol use. In contrast, drugs used in MAT identified for residual-PTU spanned a wider range of SUDs (e.g., varenicline, methadone hydrochloride, and disulfiram).

### Genetic correlations

Next, we further characterized the residual SUDs by estimating the genetic correlations of each of the four univariate (original) SUD GWAS, and the GWAS of the residual SUDs (after removing genetic influences shared with other externalizing conditions in the Externalizing model) with external correlates (Figure 5a and Supplementary Tables 20-23). Two notable patterns emerged. First, residual SUD effects appeared to capture substance specific effects in an expected manner. For example, all four univariate SUDs were significantly genetically correlated with maximum alcoholic beverage (rGs ranged from .30-.76); however, after accounting for variance shared with other externalizing traits, only residual-PAU remained significantly correlated (rG = .41, p < .001). Second, many residual SUD genetic effects retained significant associations with other forms of psychopathology, highlighting the complex associations between SUDs and both internalizing and thought disorder psychopathology. In other words, the residual genetic effects reflect not only substance specific effects like drug metabolism, but also genetic influences that impact that SUD through other mechanisms not shared with externalizing.

**Figure 5.**
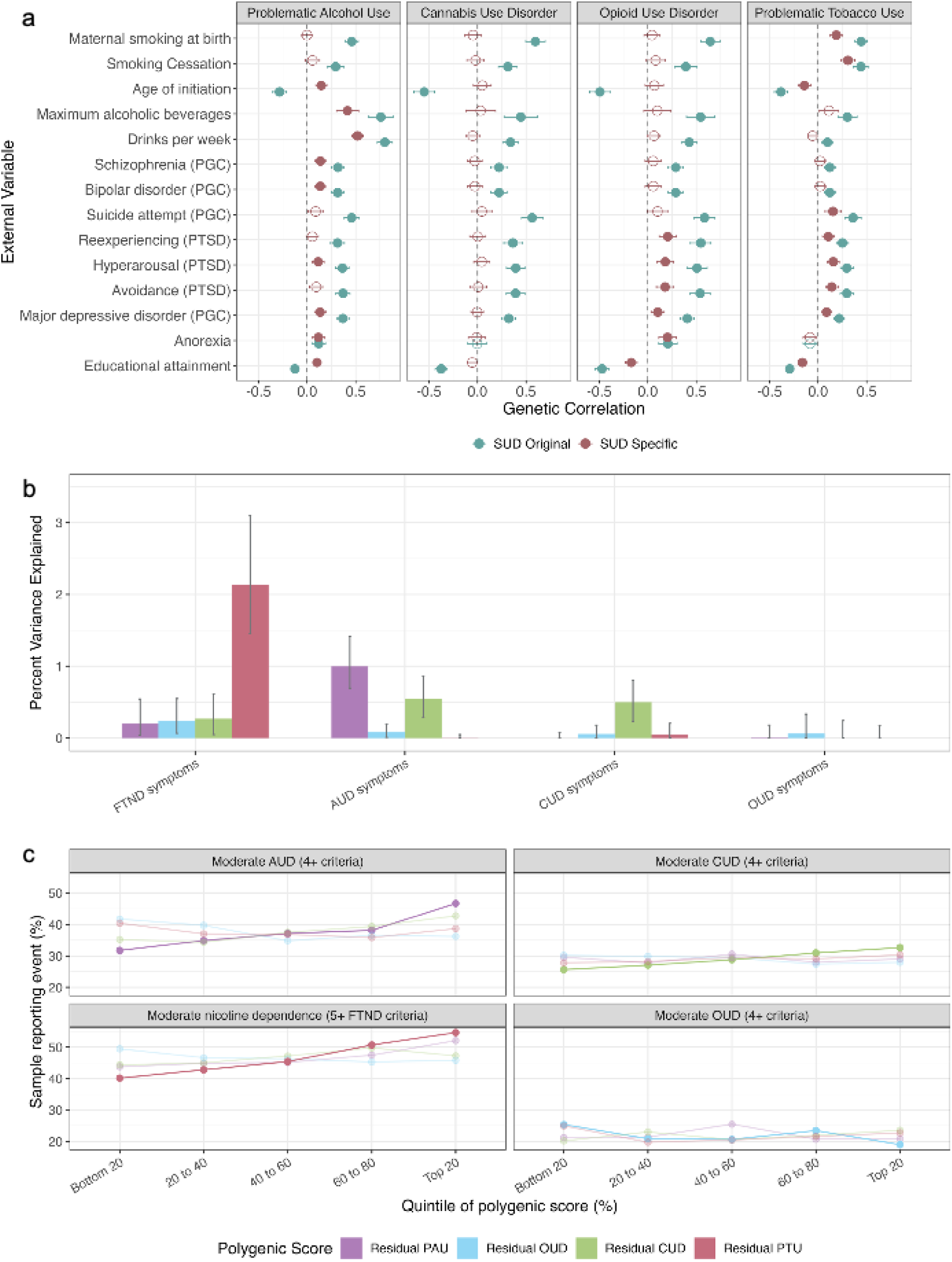
Genetic association analyses with residual SUDs. (**a**) genetic correlations and their 95% confidence intervals of the original SUD indicator GWAS (blue) and the residual SUD genetic effects (red) with relevant outcomes, (**b**) *R^2^* estimates and their 95% confidence intervals of variance explained by residual SUD PGS in SUDs in COGA and (**c)** corresponding relative risk of moderate substance use disorder across quintiles of PGS. Matched SUD phenotype and residual SUD PGS are shown in a darker color relative to unmatched PGS.

### Polygenic scores

We calculated polygenic scores from GWAS of each latent genomic factor (Externalizing, Behavioral Disinhibition, and SUD) among EUR individuals from the Collaborative Study on the Genetics of Alcoholism (COGA; *N* =7,530), a multi-site family-based study^39^ and All of Us (*N*_max_ = 77,442), a national cohort study^40,41^. We also calculated PGS for the residual SUD effects (for alcohol, opioid, tobacco, and cannabis use disorders) in COGA. These analyses allowed us to compare the total variance explained by each of the PGS and identify the specificity, or loss thereof, in polygenic prediction of SUDs. In COGA, we report associations with alcohol use disorder (AUD), cannabis use disorder (CUD), opioid use disorder (OUD), and symptoms from the Fagerstrom Test for Nicotine Dependence (FTND)^42^. In All of Us, we report associations with AUD, tobacco use disorder (TUD), and drug use disorder (DUD), which includes all other illicit substances other than alcohol and tobacco.

#### Factor PGS

EXT_PGS_ explained the most variance in all SUDs across both samples except AUD diagnosis in All of Us, which was slightly better predicted by SUD_PGS_. In COGA, EXT_PGS_ explained between 1.6% (CUD) and 3.3% (AUD) of the variance and SUD_PGS_ explained .39% (OUD) and 2.9% (FTND symptoms; Supplementary Table 24). In All of Us, EXT_PGS_ explained between 3.1% (AUD) and 7.1% (TUD) of the variance in SUD diagnosis and SUD_PGS_ accounted for 3.1% (AUD) and 4.4% (DUD; Supplementary Table 25).

#### Residual SUD PGS

We next compared the variance explained by the residual SUD PGS in SUDs in COGA (Figure 5a). Here, too, we saw evidence of substance-specific associations such that FTND, AUD, and CUD symptoms were most strongly associated with the PGS that corresponds to their substance (e.g., resPTU_PGS_ was most strongly associated with FTND symptoms). In Figure 5b, we show the rates of moderate SUD across quintiles of each residual PGS, with the matched SUD phenotype and residual SUD PGS shown in a darker color. Here we see that, especially for residual-PAU and PTU PGS, the prevalence of the disorder is lowest in the lowest quintile and increases in a more linear fashion across quintiles of risk in matched relative to unmatched PGS.

## Discussion

Despite evidence that SUDs share phenotypic^43^ and genetic^8,12,18^ variance with each other and with other externalizing traits, gene identification efforts continue to study individual SUDs in isolation or treat them as distinct from externalizing. This leaves potentially important genetic variance untagged and may, in part, explain the slow progress in gene discovery for certain SUDs relative to other complex behavioral phenotypes^44^. The role of externalizing in SUDs is also important for prevention, intervention and treatment. If SUD genetic risk largely reflects broad externalizing risk, it suggests that individuals at risk for or current experiencing an SUD are also at risk for other externalizing problems, which should be taken into account in prevention and intervention efforts. Here, we drew from the phenotypic classification and twin literature on the nature of genetic influences on SUDs^5,7,12^ and capitalized on recent advances in multivariate statistical genetic methods^16^, to model the shared genetic architecture of SUDs and other externalizing phenotypes with the goal of improving gene discovery for SUDs.

We performed multivariate GWAS, biological annotation, and genetic association analyses using the factors and residual SUD genetic effects identified in our study to address our three main questions related to improving insights into the neurobiology of broad and specific genetic effects on SUD. Our results demonstrate that modeling genetic covariance of SUDs alongside related externalizing traits improves gene discovery for SUDs and did not result in loss of specificity to detect SUD-specific effects. We base this argument on the observation that, across analyses, the broad externalizing factor was better powered to detect genetic variants for and account for variance in SUDs relative to the more specific factors and that analyzing SUDs separately did not result in novel findings related to SUDs.

Analysis of the residual SUD genetic effects revealed several interesting insights into their neurobiology. First, many genes identified for the residuals were involved in substance specific pharmacokinetic and pharmacodynamic processes (e.g., *ADH1B, CHRNA5*), potentially indicating that genetic liability to addiction is made up of risk general to externalizing and risk related to an individuals’ biological sensitivity to each specific substance. Second, genetic correlation analyses suggested that most cross substance genetic associations (e.g., correlations between CUD and alcohol phenotypes) are explained by shared externalizing variance. In addition, most SUDs retained significant associations with other forms of psychopathology (e.g., internalizing and thought disorders), highlighting the complex interface between SUDs and a broad range of psychopathology. Finally, the residual PGS showed substance specific prediction such that the residual PGS for a specific SUD best predicted that SUD in an independent sample. This highlights the potential translational utility of broad and specific PGS whereby a broader metric of risk captures an individual’s general liability to addiction whereas specific metrics can provide insight into risk for problems with specific substances. Relative to the previous externalizing and addiction risk GWAS^8,17^, we identified 187 novel genetic variants using our expanded externalizing model, including several previously implicated in substance use traits. Our expanded externalizing factor also picked up genes not identified by the other factors and which were enriched for substance use outcomes, highlighting the benefit of joint analysis for these phenotypes.

These findings should be interpreted in light of a few limitations. First and foremost, our analyses include only data from participants of European ancestry, thereby limiting the generalizability of our results. This was a practical choice as we relied on previously published GWAS and sufficiently powered GWAS of non-European ancestry samples are not yet available for these outcomes. Second, although our sensitivity analyses indicated that this phenotype did not exert undue influence on the composition of externalizing, we note that sources of individual differences in age at first sex might differ across populations^45,46^. Third, results for individual residual analyses are dependent on both the power of the original GWAS indicator and the loadings of that indicator on its factor, which determines how much variance is leftover to analyze. Finally, although SUDs manifest the strongest relationships with other externalizing phenotypes, they also have a complex interface with many other forms of psychopathology, including internalizing^47,48^, which are not modeled in the current study.

## Methods

### Multivariate GWAS

This study was approved by the Rutgers University IRB. We used Genomic SEM^16^ to estimate SNP effects in two models (Figure 1). In our two-factor model, the SUD factor included problematic alcohol use PAU)^49^, problematic tobacco use (PTU)^9^, opioid use disorder (OUD)^24^, and cannabis use disorder (CUD)^25^. The remaining non-SUD indicators from the original externalizing model (attention deficit hyperactivity disorder [ADHD]^19^, risk taking [RISK]^22^, number of sexual partners [NSEX]^22^, age at first sex [FSEX]^22^, smoking initiation [SMOK]^21^, and cannabis initiation [CANN]^20^) loaded onto a behavioral disinhibition factor (boxes B and C in Figure 1). In our common factor model, all behavioral disinhibition phenotypes and SUDs loaded onto a common externalizing factor (Box A in Figure 1). We retained the same GWAS as were used in the respective original multivariate GWAS except in the case of OUD, for which a better powered GWAS was available.

Clumping was performed in Functional Mapping and Annotation of Genome-Wide Association Studies (FUMA) version 1.6.1^50^ using an *r*^2^ threshold of ≥ 0.6 to define independent significant SNPs, a second threshold of *r*^2^≥ 0.1 to define lead SNPs, and a maximum distance between LD blocks of 250kb to merge into a locus.

To characterize differences in statistical power among the multivariate GWAS, we examine the mean χ^2^, λ_GC_, and number of genome-wide significant risk loci for each factor and residual SUD (Supplementary Table 1c). We evaluated the novelty of our findings by comparing the genomic risk loci identified in our analyses to 1) loci identified in the original externalizing and addiction risk GWAS and 2) loci previously identified for substance use phenotypes in the GWAS literature. This latter test was performed by comparing the genomic risk loci for our factors and correlated SNPs (*r*^2^ > 0.1) to those in the NHGRI-EBI GWAS Catalog^29^ (version e114_r2025-06-27). Finally, we assessed the relative performance of our two models by comparing the degree of heterogeneity of SNP effects. We did so by calculating Q_SNP_ heterogeneity statistics, which can be used to identify SNPs that have an effect on one or more indicator phenotypes that is better explained by pathways independent of the factor. In other words, the Q_SNP_ test is designed to ensure that SNPs are not being detected with the shared factor primarily due to their association with one of the constituent indicators. If a factor is truly capturing the majority of genetic variance shared among its indicator phenotypes, there will be few Q_SNP_ loci relative to the number of factor loci.

### Biological annotation

#### Gene-based methods

We used four methods to identify genes associated with the three latent genomic factors. First, we used multi-marker analysis of genomic annotation (MAGMA; version 1.08)^30^, in which genome-wide SNPs were mapped to 18,235 protein-coding genes from Ensembl v102, and SNPs within each gene were jointly tested for association with each factor. We evaluated Bonferroni corrected significance adjusted for the number of genes (one sided p < 2.74×10^-6^). Second, we used Hi-C-coupled MAGMA (H-MAGMA; version XX)^33^, which builds on MAGMA by leveraging chromatin interaction profiles from human brain tissue. We evaluated Bonferroni corrected two-sided p-value thresholds, adjusted for multiple testing within each analysis (two-sided ps < 1.72×10^-7^ for residual-PAU and 1.62×10^-7^ for all other phenotypes). Next, we used MetaXcan^31^ to conduct a Transcriptome-Wide Association Study (TWAS) using genetically regulated expression models from GTEx v8^51^. This analysis leveraged GWAS summary statistics to estimate gene-trait associations. Within-tissue Bonferroni correction was applied to identify statistically significant TWAS genes. Finally, we used summary-data-based Mendelian randomization (SMR)^32^ to 1) test the extent to which gene expression mediated the relationship between SNPs and the phenotype and 2) identify genes that are more likely to be functionally relevant to the phenotype using the heterogeneity in dependent instruments (HEIDI) test. The HEIDI test distinguishes causality and pleiotropy models, in which the effect of a genetic variant on a trait is mediated by gene expression or where the genetic variant has direct effects on both the trait and expression, from the linkage model, in which associations between gene expression and trait are due to LD between two distinct causal variants. Evidence of causal or pleiotropic effects suggests that the gene should be prioritized for follow up analyses. We identified genes of interest as those that met SMR test Bonferroni correction significance threshold and had a HEIDI test two-sided p-value > .05. We also used MAGMA to identify genes associated with each residual SUD phenotype.

#### Network Analysis

We employed gene-network analysis to compare the biological processes underlying Externalizing, Behavioral Disinhibition, and SUDs. Using a random-walk^34^ algorithm within the PCNet2.0^35^ interactome, we generated gene-networks for each latent genomic factor. As described previously^17,52^, network propagation was computed using NetColoc^34^, using an alpha=0.5, and a z-score cutoff of 3. We generated a multiscale systems map of the for each network using the Hierarchical community Decoding Framework (HiDeF v1.1 Beta) algorithm as implemented in Cytoscape’s Python package CDAPS^53^, using a maximum resolution of 2, and enrichment for GO terms limited to terms with 50-1,000 annotations, a minimum of 3 genes overlapping, and enrichment two sided p-value<0.05. All significant annotations for each community were manually aggregated into a single label for each community. GWAS Catalog enrichment was calculated using Experimental Factor Ontology based phenotype grouping for the GWAS catalog, downloaded from https://www.ebi.ac.uk/gwas/docs/file-downloads on 7 August 2024. Enrichment was calculated via hypergeometric test against all PCNet2.0 nodes, and Bonferroni corrected for all traits in the Experimental Factor Ontology.

#### Drug Repurposing Analyses

Factor genes identified by at least three methods and residual genes identified in at least one method were mapped to the ^54^. We queried broadly for all FDA approved drugs as well as looked specifically for following FDA approved drugs used in the medication assisted treatment (MAT) of any substance use disorder: buprenorphine, naltrexone, naloxone hydrochloride, methadone hydrochloride, acamprosate calcium, disulfiram, varenicline, bupropion hydrochloride, and baclofen.

### Genetic Correlations

Genetic correlations of the univariate (i.e., original) SUDs and residual SUDs with 39 external correlates were estimated in Genomic SEM. External correlates were chosen based on their putative relevance to SUDs and included variables from the domains of cognitive traits, other forms of psychopathology, personality, and substance use. Residual correlations were estimated from the Externalizing model, in which loadings of SUDs on the common Externalizing factor and their genetic correlation with an external correlate were simultaneously estimated. Statistical significance was determined using a Bonferroni correction.

### Polygenic scores

#### Target samples

We calculated polygenic scores among EUR individuals from the Collaborative Study on the Genetics of Alcoholism (COGA; *N* = 7,530) and All of Us (*N*_max_ = 77,442).

#### COGA

The Collaborative Study on the Genetics of Alcoholism is a large, multi-site, family-based study incorporating multiple generations with the goal of studying etiological influences on alcohol use disorders (AUDs)^39^. Individuals in treatment for alcohol use disorder and their families were recruited into the study along with a smaller number of community-based comparison families beginning in 1991. Data were collected over four waves, culminating in data available for those included in the initial assessment, follow-up of individuals in the first wave, a prospective wave, which focused on longitudinal assessments of youth in case and comparison families, and an ongoing wave of participants now in midlife and later life stages. The present study includes data from individuals in waves 1-3 who were of EUR ancestry and had GWAS data available.

DNA samples were genotyped using the Illumina Human1 M array (Illumina, San Diego, CA), The Illumina Human OmniExpress 12V1 array (Illumina), the Illumina 2.5M array (Illumina) or the Smokescreen genotyping array (Biorealm LLC, Walnut, CA)^55^. Data were imputed to 1000 Genomes (Phase 3) and SNPs with a minor allele frequency < .01, were genotyped at a rate < .95, or that violated Hardy-Weinberg equilibrium were excluded. Principal components (PCs) were calculated using Eigenstrat^56^ and 1000 Genomes (Phase 3, version 5). The first ten PCs, age, and sex were used as covariates in the polygenic score analyses (PGS). Additional details about genotyping and quality control procedures have been published elsewhere^57,58^. We report associations with problematic substance use (symptoms of AUD, cannabis use disorder [CUD], opioid use disorder [OUD], and other SUD as well as the Fagerstrom Test for Nicotine Dependence^42^).

#### All of Us

All of Us is an observational cohort study of diverse, adult (age ≥ 18) participants, intended to be representative of the United States with the goal of studying the effects of lifestyle, environment, and genomics to improve health outcomes^40,41,59^. Data collection began in May 2018 and is ongoing. Participants are recruited through health care organizations, other community enrollment sites, or by enrolling directly online. Participants give informed consent and complete a basic health survey online and are then invited to undergo a physical examination and to provide a biospecimen at an affiliated health care site. If consent is provided, additional information is collected by linking participants’ electronic health records (EHR) and by completing periodic surveys assessing lifestyle, wellbeing, physical health, and behavior. Participants in release 7 (May 6, 2018 to February 23, 2023) with linked EHR and whole genome sequencing data were included in the current study.

Participants were genotyped using the Illumina Global Diversity Array. The All of Us Research Program completed genetic ancestry classification analyses using a random forest classifier to predict ancestry from principal components (PCs). PCs, age, and sex were used as covariates in the PGS analyses. Outcome SUDs include AUD, TUD, and other drug disorder (i.e., use disorder for all other drugs) present in the EHR. Diagnoses for all disorders were based on phecodes, which are clusters of related billing codes from the International Statistical Classification of Diseases 9^th^ and 10^th^ revisions. Individuals with two or more phecodes related to a single disorders were categorized as cases.

#### Calculation of PGS

We used PRS-CS^60^ to adjust original GWAS beta weights for linkage disequilibrium and Plink2^61^ to construct each PGS from these weights. We evaluated the incremental R^2^/pseudo-R^2^ (Δ*R*^2^) attained by adding the polygenic score to a regression with baseline covariates (e.g., age, sex, and ancestry PCs). We used least squares regression for continuous outcomes and logistic regression for categorical ones and adjusted the standard errors in COGA to account for the family structure. Regression models were estimated using the *estimatr* package in R^62^. We estimated 95% CIs for Δ*R*^2^ using bootstrapping (1,000 iterations) in the *boot* R package ^63^.

## Acknowledgements

This work was supported by the National Institute on Alcohol Abuse and Alcoholism (R01AA015416 to DMD, K02AA018755 to DMD, U10AA008401 to DMD, P50AA022537 to DMD, T32AA028254 to HEP), the National Institute on Drug Abuse (K01DA059657 to HEP, R01DA050721 to DMD, DP1DA054394 to SSR), the Tobacco-Related Disease Research Program (T32IR5226 to SSR and 28IR-0070C to AAP), and the National Institute of General Medical Sciences (T32 GM008666 to BSL).

The authors thank **The Externalizing Consortium.** Principal Investigators: Danielle M. Dick, Philipp Koellinger, K. Paige Harden, Abraham A. Palmer. Lead Analysts: Richard Karlsson Linnér, Travis T. Mallard, Peter B. Barr, Sandra Sanchez-Roige. Significant Contributors: Irwin D. Waldman. The Externalizing Consortium has been supported by the National Institute on Alcohol Abuse and Alcoholism (R01AA015416 – administrative supplement to DMD), and the National Institute on Drug Abuse (R01DA050721 to DMD). Additional funding for investigator effort has been provided by K02AA018755, U10AA008401, P50AA022537 to DMD, as well as a European Research Council Consolidator Grant (647648 EdGe to Koellinger). The content is solely the responsibility of the authors and does not necessarily represent the official views of the above funding bodies. The Externalizing Consortium would like to thank the following groups for making the research possible: **23andMe Inc, Add Health, Vanderbilt University Medical Center’s BioVU, Collaborative Study on the Genetics of Alcoholism (COGA), the Psychiatric Genomics Consortium’s (PGC) Substance Use Disorders working group, UK10K Consortium, UK Biobank,** and **Philadelphia Neurodevelopmental Cohort.** All code necessary to replicate this study is available upon request.

The authors thank **Million Veteran Program (MVP)** staff, researchers, and volunteers, who have contributed to MVP, and especially participants who previously served their country in the military and now generously agreed to enroll in the study.

We gratefully acknowledge ***All of Us*** participants for their contributions, without whom this research would not have been possible. We also thank the National Institutes of Health’s *All of Us* Research Program for making available the participant data examined in this study.

Finally, we thank **The Collaborative Study on the Genetics of Alcoholism (COGA)**, Principal Investigators B. Porjesz, V. Hesselbrock, A. Agrawal; Scientific Director, A. Agrawal; Translational Director, D. Dick, includes ten different centers: University of Connecticut (V. Hesselbrock); Indiana University (H.J. Edenberg, T. Foroud, Y. Liu, M.H. Plawecki); University of Iowa Carver College of Medicine (S. Kuperman, J. Kramer); SUNY Downstate Health Sciences University (B. Porjesz, J. Meyers, C. Kamarajan, A. Pandey); Washington University in St. Louis (L. Bierut, A. Agrawal, S. Hartz); University of California at San Diego (M. Schuckit); Rutgers University (J. Tischfield, D. Dick, R. Hart, J. Salvatore); The Children’s Hospital of Philadelphia, University of Pennsylvania (L. Almasy); Icahn School of Medicine at Mount Sinai (A. Goate, P. Slesinger); and Howard University (D. Scott). Other COGA collaborators include: C. Holzhauer, M. Hesselbrock (University of Connecticut); J. Nurnberger Jr., L. Wetherill, X., Xuei, D. Lai, S. O’Connor, (Indiana University); G. Chan (University of Iowa; University of Connecticut); D.B. Chorlian, J. Zhang, P. Barr, S. Kinreich, G. Pandey, Z. Neale (SUNY Downstate); N. Mullins (Icahn School of Medicine at Mount Sinai); A. Anokhin, K. Bucholz, F. Dong, A. Hatoum, E. Johnson, V. McCutcheon, J. Rice, S. Saccone (Washington University); F. Aliev, Z. Pang, S. Kuo, S. Brislin (Rutgers University); A. Merikangas (The Children’s Hospital of Philadelphia and University of Pennsylvania); H. Chin and A. Parsian are the NIAAA Staff Collaborators. We continue to be inspired by our memories of Henri Begleiter and Theodore Reich, founding PI and Co-PI of COGA, and also owe a debt of gratitude to other past organizers of COGA, including Ting- Kai Li, P. Michael Conneally, Raymond Crowe, and Wendy Reich, for their critical contributions. This national collaborative study is supported by NIH Grant U10AA008401 from the National Institute on Alcohol Abuse and Alcoholism (NIAAA) and the National Institute on Drug Abuse (NIDA).

## COGA Collaborators

Bernice Porjesz, Victor Hesselbrock, Tatiana M. Foroud, Arpana Agrawal, Howard J. Edenberg, John I. Nurnberger Jr, Yunlong Liu, Samuel Kuperman, John Kramer, Jacquelyn L. Meyer, Chella Kamarajan, Ashwini K. Pandey, Laura Bierut, John Rice, Kathleen K. Bucholz, Marc A. Schuckit, Jay Tischfield, Andrew Brooks, Ronald P. Hart, Laura Almasy, Danielle M. Dick, Jessica E. Salvatore, Allison Goate, Manav Kapoor, Paul Slesinger, Denise M. Scott, Lance Bauer, Leah Wetherill, Xiaoling Xuei, Dongbing Lai, Sean J. O’Connor, Martin H. Plawecki, Spencer Lourens, Laura Acion, Grace Chan, David B. Chorlian, Jian Zhang, Sivan Kinreich, Gayathri Pandey, Michael J. Chao, Andrey P. Anokhin, Vivia V. McCutcheon, Scott Saccone, Fazil Aliev, Peter B. Barr, Hemin Chin & Abbas Parsian

## Data Availability

The summary statistics used for the current study are from previously published studies and no new data were collected. These studies are described in the text. Some of these studies include data which have restricted access to protect the privacy of the study participants. The full sets of externalizing and residual SUD summary statistics generated by this study can be made available to qualified investigators who enter into an agreement with 23andMe that protects participant confidentiality and who have access to MVP data. Once these requests have been satisfied, investigators may request to use our summary statistics via the procedures detailed at https://externalizing.rutgers.edu/request-data/.

## Author Contributions

HEP, DMD, and PBB contributed to the conception and design. HEP, CC, BL, JG, and PB were responsible for analysis of the data. All authors contributed to the interpretation of the data. HEP and BSL were responsible for drafting the original manuscript and all authors contributed to revising and editing the manuscript. PBB and DMD were responsible for supervision of this work.

