## Supplementary Text for "Multivariate genetic of 2.2 million individuals demonstrate genetic influences on substance use disorders operate via behavioral disinhibition and substance-specific risk"

**Supplementary Results & Figures**

Herein, we report additional details about analyses included in the main text as well as analyses that are relevant to our research questions but were not included in the main text due to space constraints.

**Multivariate GWAS**

We identified 708 loci in the single factor Externalizing model, in which SUDs were modeled as part of the externalizing spectrum. In the two-factor model, we identified 631 and 48 genomic risk loci (679 total) for the Behavioral Disinhibition and SUD factors, respectively (Figure 2; Supplementary Table 3). Although we identified the greatest number of risk loci using the broad Externalizing factor, we observed an increased number of hits for Behavioral Disinhibition and SUD relative to their original multivariate GWAS as well. Of the 631 Behavioral Disinhibition loci and their correlates, 94 (15%) were not identified in the previous Externalizing GWAS. Similarly, of the 48 SUD loci and their correlates, 33 (69%) were not identified in the previous Addiction Risk GWAS, and 6 (13%) were not previously associated with any substance use trait in the GWAS Catalog. These analyses suggest that even when modeled as separate but correlated dimensions, the addition of other externalizing traits boosts power to identify genes involved in SUDs.

We next compared the degree of overlap between loci and their correlates identified across the three factors. Only one locus was identified for all factors, rs34488670, which is mapped to *SEMA6D*, and has been associated with a variety of risky taking behaviors and age of smoking initiation as reported in the GWAS Catalog. Of the 708 loci and their correlates for Externalizing, 272 were also identified for Behavioral Disinhibition, 2 were identified for SUDs, and 135 were unique to Externalizing. The loci for Behavioral Disinhibition were mostly shared with Externalizing (546), but 64 were unique to that factor and none were shared with SUDs but not Externalizing. Finally, most loci for SUDs were shared across all factors or with Externalizing (27 total), but 21 were unique to SUDs.

Finally, we performed SNP-level tests of heterogeneity (Q_SNP_) to investigate the extent to which SNPs identified with the factors had consistent, pleiotropic effects on the constituent indicators of that factor. Significant Q_SNPs_ indicate variants that exert heterogeneous effects on the factor indicators, rather than reflecting shared effects on the whole factor. We identified 134 Q_SNPs_ for the Externalizing model and 112 Q_SNPs_ for the two-factor Behavioral Disinhibition and SUD model, suggesting that SNP effects in the common factor model were not substantially more heterogeneous than those in the two-factor model. Only one Q_SNP_ loci from the Externalizing model overlapped with any of the 708 significant genomic risk loci identified for that factor. This was rs4702, a noncoding transcript variant located in the gene *FURIN* that has previously been associated with OUD^28^. Similarly, only three significant loci from the Behavioral Disinhibition and SUD model were Q_SNP_ loci. These loci were rs4702 and rs1229984, a missense variant in *ADH1B* associated with alcohol phenotypes^29,30^, and rs13135092, an intronic variant located in *SLC39A8* also previously associated with AUD^24^. Of the 38 loci identified across all four residual SUDs, 14 overlapped with Q_SNP_ loci from the Externalizing and/or SUD factor, suggesting that Q_SNP_ loci represent substance-specific genetic effects.

**Tissue Expression**

We also used MAGMA tissue expression analysis to test the relationship between expressed genes in different tissues across the three factors and three of the four residual SUDs (analyses were not performed for residual-OUD as no significant SNPs were identified in this analysis; Supplementary Figure 2-3). Tissue expression analysis used weights from GTEx v8^44^. We also directly compared the standardized beta coefficients of associations for 15 brain, liver, and lung tissues to highlight any differences in direction or magnitude of associations across the three factors (Supplementary Figure 4) and found little evidence of differential associations. The direction of effects was the same across factors, but the magnitude of the associations for SUDs was significantly smaller than those for Behavioral Disinhibition and Externalizing, highlighting the differences in power, but not evidence of differences in magnitude of association.

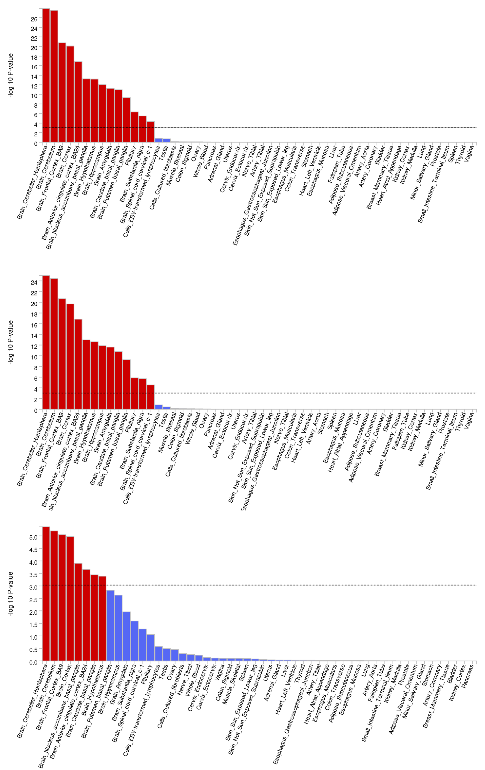

**Supplementary Figure 1.** MAGMA tissue expression with GTEx v8 tissue types, ordered by p-value for Externalizing (top), Behavioral Disinhibition (middle), and SUDs (bottom).

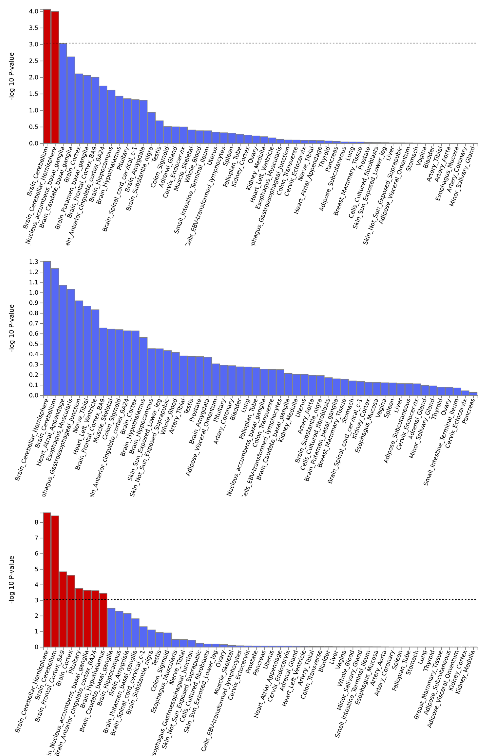

**Supplementary Figure 2.** MAGMA tissue expression with GTEx v8 tissue types, ordered by p-value for residual Problematic Alcohol Use (top), Cannabis Use Disorder (middle), and Problematic Tobacco Use (bottom).

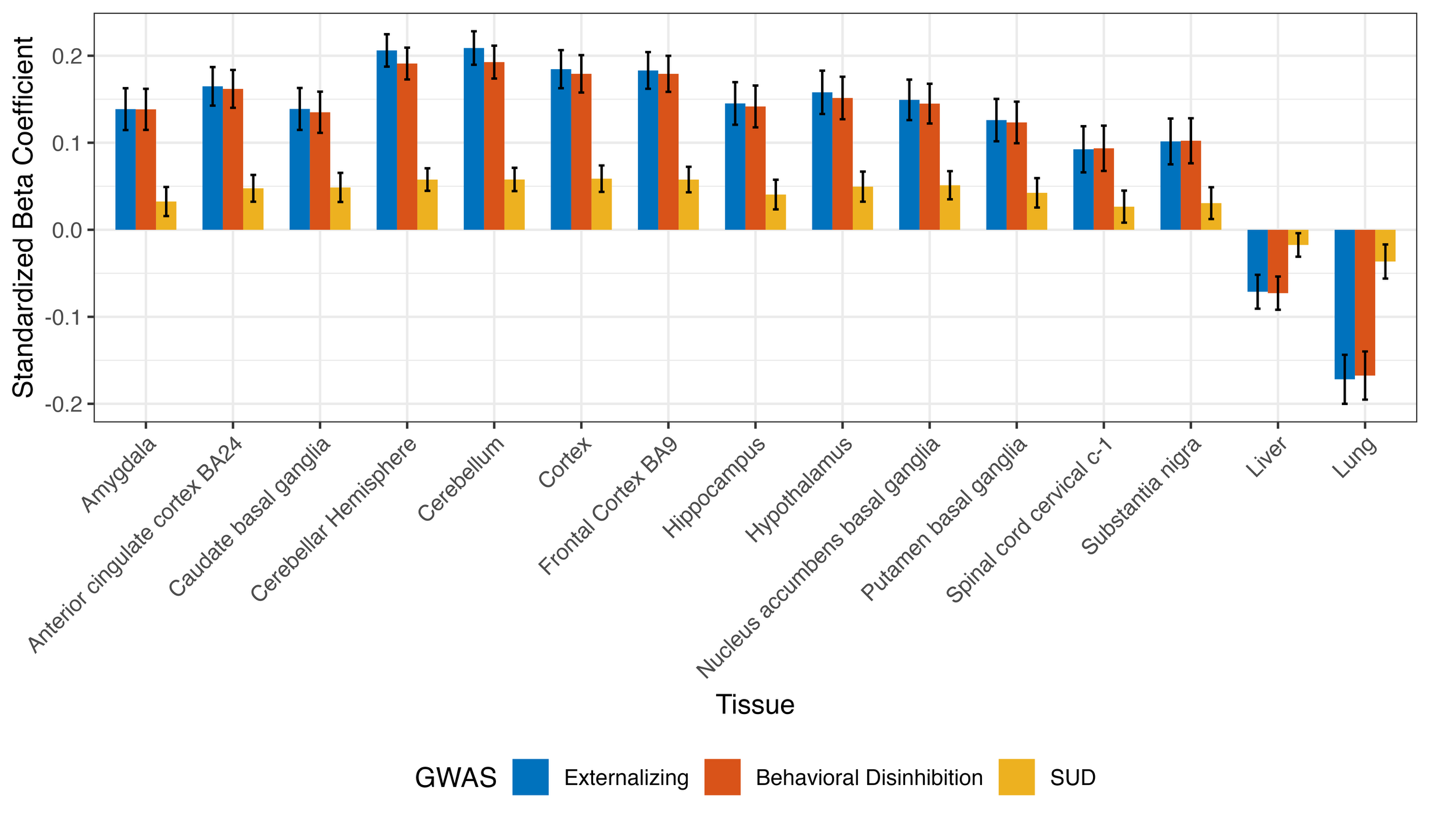

**Supplementary Figure 3.** Comparison of the standardized beta coefficients of associations for brain, liver, and lung tissues from MAGMA tissue expression analyses across the Externalizing, Behavioral Disinhibition, and SUD factors.

**Network Analysis**

We compare the biological processes underlying Externalizing, Behavioral Disinhibition, and SUDs using gene-network analysis. At the network level, the Externalizing and Behavioral Disinhibition networks showed high similarity, unlike the SUD network. The Externalizing and Behavioral Disinhibition networks shared 241 of their genes, 74 of which were present in both initial gene sets. In comparison, the SUD network shared only 5 genes with the networks for the two latent factors (Figure 4a). This is likely due to the limited power of the SUD gene set, leading to the network containing more false-positives. The Externalizing and Behavioral Disinhibition networks contained similar communities, with those for Behavioral Disinhibition similarly functioning in intracellular transport, protein oxidation and degradation, and synaptic vesicle cycle (Supplemental Figure 5a), while the SUD network communities were enriched for chromatin regulation and general biological regulation (Supplemental Figure 5b). With respect to annotation in the GWAS catalog, Externalizing was enriched for genes in psychiatric disorders (p=2.2x10^-7^), substance-related disorders (p=2.5x10^-10^), and substance abuse (p=4.5x10^-14^), and the Behavioral Disinhibition network similarly was enriched for substance-related disorders (p=5.3x10^-13^) and substance abuse (p=2.5x10^-16^). The SUD network was not significantly enriched for genes identified in any traits in the GWAS catalog. The Externalizing enrichment has highest in the cytoskeletal transport communities (Figure 4b), while the Behavioral Disinhibition was centralized on protein oxidation and synaptic vesicle cycle (Supplemental Figure 5a).

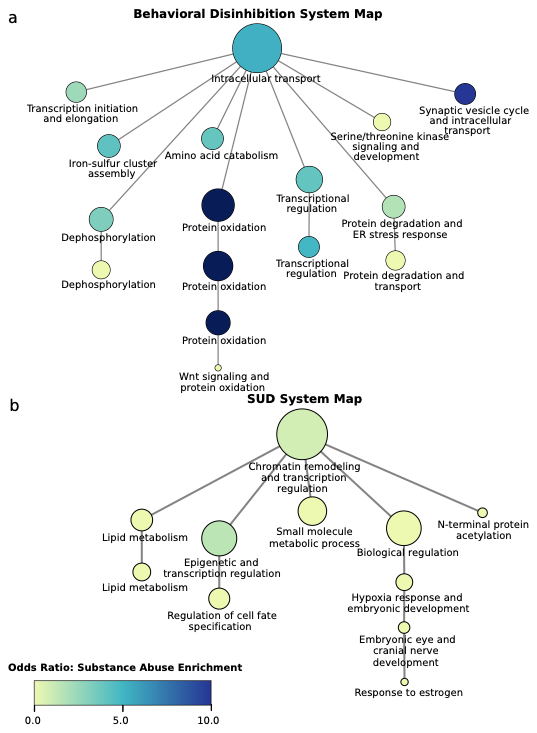

**Supplementary Figure 4**: Network analysis of Behavioral Disinhibition and SUD. System map for (a) Behavioral Disinhibition and (b) SUD networks. Community labels represent aggregated significant gene ontology enrichment labels, and node size indicates log-transformed community size, measured by number of genes. Color indicates the odds ratio of the enrichment of that community for genes annotated in the GWAS catalog for substance abuse. Only labelled communities, or those connecting labeled communities are shown

**Sensitivity Analysis with Age at First Sex**

To assess the influence of the Age at First Sex (FSEX) indicator on our results, we conducted a series of sensitivity analyses excluding this indicator from the model of EXT and compared the results to those from the full model. We first estimated the genetic correlation between the full and reduced model was 0.99 (SE = .03) and the correlation between the effect sizes from the two sets of summary statistics was .96 (SE = .001). Next, we estimated the genetic correlations between the reduced EXT model and each of the external correlates used in the main analyses (Supplementary Table 23). We then estimated the correlation between the effect sizes of the full and reduced EXT with the external correlates and found that their effect sizes were correlated at .99 (.01). This indicates that the specificity of the EXT model is relatively robust to the inclusion or exclusion of the FSEX indicator. In contrast, removing FSEX resulted in a loss of GWAS signal, as indicated by the reduced mean χ^2^ (3.21 in the reduced model vs. 3.45 in the full model), λ_GC_ (2.46 in the reduced vs. 2.56 in the full model), and genome-wide significant risk loci (651 in the reduced vs. 708 in the full model). From this we conclude that the inclusion of FSEX does not exert undue influence on the composition of the EXT factor, but does result in loss of signal, leading us to conclude that the indicator’s inclusion is warranted.
